# Differential modulation of attentional ERPs in smoked and insufflated cocaine-dependent associated with neuropsychological performance

**DOI:** 10.1101/2023.07.07.23292384

**Authors:** Agustina Aragón-Daud, Sofía Milagros Oberti de Luca, Sofía Schurmann Vignaga, Pilar Prado, Rosario Figueras, Lucia Lizaso, María Luz González-Gadea, Facundo Manes, Marcelo Cetkovich, Carla Pallavicini, Teresa Torralva, Laura Alethia de la Fuente

## Abstract

**Background:** Cocaine consumption is associated with reduced attentional event-related potentials (ERPs), namely P3a and P3b, indicating bottom-up and top-down deficits respectively. At cognitive level, these impairments are larger for faster routes of administration (e.g., smoked cocaine [SC]) than slower routes (e.g., insufflated cocaine [IC]). Here we assess these ERPs considering the route of cocaine administration. We hypothesized that SC dependent (SCD) would exhibit reduced modulation of the P3a, while both SCD and IC dependent (ICD) would show reduced modulation of the P3b.

**Methods:** We examined 25 SCD, 22 ICD matched by poly-consumption profiles, and 25 controls matched by demographic variables. We combined EEG data from the Global-Local task with behavioral data from attentional cognitive tasks.

**Results:** At the behavioral level, SCD exhibited attentional deficits in both bottom-up and top-down processes, while ICD only showed a tendency for top-down deficits. Modulation of P3a and P3b was lower in consumer groups. We observed subtle route-based differences, with larger differences in the P3a for SCD and in the P3b for ICD. Neurophysiological and behavioral data converged, with the P3a associated to bottom-up performance and P3b to top-down.

**Conclusions:** Different routes of administration lead to distinct modulations of attentional neurocognitive profiles. Specifically, SCD showed greater attentional impairment, mainly at bottom-up/P3a, while ICD showed a trend of top-down/P3b deficits. These findings emphasize the crucial role of considering the route of administration in both clinical and research settings and support the use of attentional ERPs as valid measures for assessing attentional deficits in substance abuse.

## 1. Introduction

Smoked cocaine (SC) represents a major health risk among vulnerable populations in Latin America (Castaño, 2000; Castilla et al., 2020; PJCBA, 2016). In particular, SC has a higher potential for abuse than other slower routes of administration, such as insufflated cocaine (IC), as the former facilitates reinforcement and induces neuroplasticity changes in the reward system more easily (Allain et al., 2015; Oliveira et al., 2018; Samaha and Robinson, 2005). Our previous research has shown that SC dependent individuals (SCD) exhibit greater attentional-executive impairments in cognitive tasks than IC dependent (ICD) (de la Fuente et al., 2021). However, the neurophysiological underpinnings of these deficits remain unexplored. In this work we measured attentional event-related potentials (ERPs), biomarkers of addiction and relapse (Anderson et al., 2011; Bauer, 1997; Habelt et al., 2020; Hajcak et al., 2019; Harper et al., 2021), on consumers who differed only in the route of administration. By employing state-of-the-art attentional ERPs from oddball paradigms and a comprehensive attentional neuropsychological assessment, we aimed to better understand the neural correlates of these cognitive deficits.

Oddball paradigms elicit the mismatch negativity (MMN), P3a and P3b components, reflecting pre-attentional, bottom-up, and top-down attentional processes, respectively (Conroy and Polich, 2007; Gooding et al., 2008; Hsieh et al., 2021). The MMN and P3a potentials are evoked by infrequent and salient unexpected stimuli, while the P3b is elicited by infrequent expected stimuli (Carrasco, 2011; Desimone and Duncan, 1995; Friedman et al., 2001; Näätänen et al., 2007; Polich, 2007). Studies have consistently shown bottom-up and top-down deficits signed by reduced P3a and P3b in consumers of stimulants (Iwanami et al., 1998) and other drugs (Harper et al., 2021, 2020; Houston and Schlienz, 2018; Jurado-Barba et al., 2020). Interestingly, enhanced P3a and P3b responses are elicited towards drug-related cues, suggesting attentional bias and lack of inhibition, respectively (Biggins et al., 1997; Campanella, 2021). Notably, while overall cocaine consumers exhibit executive-flexibility deficits at the cognitive level, only SCD show attentional impairments (de la Fuente et al., 2021). Considering that factors such as drug-use level and drug type can impact the P3a and P3b (Polich and Criado, 2006), these cognitive profile differences suggest the possibility of distinct modulation of these ERPs based on different routes of administration.

We hypothesized that SCD would exhibit reduced modulation of the P3a, while both SCD and ICD would show reduced modulation of the P3b. The impact of different routes of cocaine administration on these attentional ERPs remains unclear, as most studies have not accounted for this relevant variable (Anderson et al., 2011; Campanella et al., 2019; Habelt et al., 2020; Wakim et al., 2021). Given the clinical relevance of these attentional ERPs in predicting addiction and relapse (Anderson et al., 2011; Bauer, 1997; Habelt et al., 2020; Hajcak et al., 2019; Harper et al., 2021) and the distinct cognitive profiles associated with the route of cocaine administration, this study examines the differences in the P3a and P3b potential between SCD and ICD, and their relationship with neuropsychological performance.

## 2. Materials and methods

### 2.1 Participants

The study included 72 subjects, 25 SCD, 22 ICD, and 25 controls (CTR) matched based on demographic variables (Table 1). Consumers fulfilled the DSM-IV criteria for cocaine abuse disorder and were classified into groups based on the drug that motivated their hospitalization and expert consensus of three addiction psychiatrists (more information in Online Resources 1). Consumers were additionally matched by poly-consumption pattern, as measured by the adapted version of the ASSIST (World Health Organization, 2002) and MINI-Plus Interview (Sheehan et al., 1998) (Detailed information can be found in Online Resources, Table 1), and ensuring that the only significant difference between them was their preferred route of cocaine administration (*p* < .05). Both the SCD and ICD groups consumed various drugs (*p* > .05), including IC (Online Resources 1, Table 2). Controls were recruited as volunteers who did not have a history of drug abuse. Participants had no major psychiatric or neurological disorders or a high incidence of familial psychopathology (Online Resources 1, Table 3). Participants taking psychiatric medication were included only if the prescribed doses were below measurable at EEG (Macoveanu, 2014) (Online Resources 1, Table 4). All participants provided written informed consent, and the study was approved by the ethics committee at Favaloro University (N◦ 609/16, record 554).

**Table 1.** Demography.

|  | CTR | SCD | ICD | Statistics |
| --- | --- | --- | --- | --- |
|  | (N = 25) | (N = 25) | (N = 22) |  |
| Age* | 19.60 (2.62) | 20.04 (2.28) | 20.64 (2.96) | $F_{(2, 72)} = 0.91, p = .40$ |
| Education* | 9.47 (1.94) | 8.76 (1.83) | 9.45 (1.56) | $F_{(2, 72)} = 2.03, p = .14$ |
| Biological sex (M:F) | 22:3 | 23:2 | 21:1 | $X^2 = 0.88, p = .64$ |
| Handedness (R:L) | 25:0 | 24:1 | 22:0 | $X^2 = 1.91, p = .38$ |
| ESOMAR<br>(a:b:ca:cb:d:e) † | 0:0:1:1:15:7 | 0:1:1:3:9:6 | 0:1:5:4:6:1 | $X^2 = 15.46, p = .05$ |
| NB (URB:SBN:Street)<br>†† | 11:13:0 | 5: 11:1 | 13: 5:0 | $X^2 = 8.45, p = .08$ |

\*Values are expressed as mean (standard deviation). † The European Society for Opinion and Marketing Research (ESOMAR) for socio- economical capacity categories are very-high (a), high (b), middle-high (ca), middle (cb), middle-low (d), and low (e). †† unsatisfied basic needs (UBN), satisfied basic needs (SBN), or living in the street. CTR: Control; SCD: Smoked cocaine-dependent; ICD: cocaine hydrochloride -dependent

**Table 2.** Factorial analysis.

| NPS Variable | F1: Bottom-up | F2: Top-down |
| --- | --- | --- |
| Digit forward | 0.68 | 0.34 |
| Trail 1 (RAVLT) | 0.40 |  |
| TMT A* |  | 0.54 |
| TMT B* |  | 0.43 |
| Corsi forward |  | 0.55 |
| LNST | 0.31 | 0.54 |
| Symbol-Digit |  | 0.31 |
| Cumulative variance explained | 31% | 18% |
| Hypothesis test for 2 Factors: Chi-square (8) = 1.76 , $p = 0.98$ | | |

\*Variables inverted before factor analysis; TMT A = Trail Making A; TMT B = Trail Making B; LNST = Letter-Number Sequencing Test; RAVLT = Rey- Auditory Verbal Learning Test.

### 2.2 Attentional cognitive tasks

Participants completed a neuropsychological assessment focused on the attentional domain. The battery included Forward and Backwards Digit span (Weschler, 1999), Forward and Backwards Corsi block-tapping test (Corsi, 1972), Symbol-Digit Modality (Smith, 1973), Trail Making Test (Part A and B) (Reitan and Wolfson, 1993), Letter and Number Sequencing Test (LNST) (Weschler, 1999), Stroop test (Stroop, 1935), and the Trail 1 and Distractor List of the Rey-Auditory Verbal Learning Test (RAVLT) (Rey, 1941). A detailed description of the tests and scoring can be found in the Online Resources 2.

### 2.3 Online attentional task during EEG

We used an adapted version (Chennu et al., 2013) of the Global Local Task (Bekinschtein et al., 2009) while recording high-density EEG (hd-EEG) signals. The task, illustrated in Fig 2A., was composed of sequences of 5 complex tones of different frequency (type A or B). The first four tones were always monaural and of one type (either A or B), while the fifth tone could either be identical or different. If all five tones were equal, the sequence was classified as a local standard condition (AAAAA or BBBBB). If the fifth tone was different, the sequence was classified as a local deviant condition (AAAAB or BBBBA). The sequences were grouped into blocks that established a global regularity: one sequence (either local standard or deviant) accounted for 71.5% of the sequences in that block and constituted a global standard condition. While 14.25% violated this global regularity established across seconds and constituted a global deviant condition. The remaining 14.25% were global standard but interaural, which was not analyzed in this study. Participants were instructed to pay attention and report the number of global deviations, which elicit the P3b. Local deviation conditions elicit the MMN and P3a. For a detailed explanation, refer to Online Resources 3.1.

### 2.4 High-Density EEG data collection and preprocessing

Biosemi Active-two 128-channel system were used to record hd-EEG signals in a conditioned room (acoustically and electrically insulated). The signals were preprocessed using the EEGLAB Toolbox from Matlab (version 2014a), including resampling (256 Hz), band pass filtering (0.5 to 25 Hz), re-referencing (average), and interpolation (spherical method), for details see Online Resources 3. EEG epochs were baseline corrected and noisy epochs were rejected using an automated procedure (Zich et al., 2015) and confirmed by visual inspection. We eliminated ocular artifacts using independent component analysis and generated grand-averages for each group by condition. For a detailed explanation, refer to Online Resources 3.2.

### 2.5 Statistical analysis

#### 2.5.1 Attentional cognitive tasks

To compare performance in individual cognitive tests and factor scores between groups we used non-parametric Kruskal-Wallis tests with Wilcoxon-Mann-Whitney tests for post hoc comparisons, corrected by False Discovery Rate (FDR) with a significance level given by *p*-FDR corrected <.05. Further analysis was restricted to cognitive test with significant differences between consumers and CTR. To identify bottom-up and top-down processes from attention performance, we assessed theoretical contributions, modular structure, and factorial analyses. To construct the graph (Fig. 1B), each cognitive task was represented by nodes connected them with weights based on the non-parametric Spearman correlation between performance vectors across subjects. The resulting networks were visualized using the ForceAtlas 2 layout in Gephi (Bastian et al., 2009) (https://gephi.org/). ForceAtlas 2 is a two-dimensional representation of the network, where link weights (Spearman correlations) are modeled as springs and nodes as point charges of the same sign. Attraction and repulsion are calculated using Hooke’s law (Hooke, 1678) and Coulomb’s law (Coulomb, 1785). For the modularity analysis we used the Louvain agglomerative algorithm (Blondel et al., 2008) with a resolution parameter γ=1 to identify sub-networks with dense internal connections and sparse external connections (i.e. modules). In order to compress the information from the multiple cognitive task into two factors reflecting bottom-up and top-down processes, we used a factorial model based on Varimax rotation (Wassing et al., 2016) and assessed its consistency with the theoretical and modular structure. Individual scores for two main factors in an Exploratory Factorial Analysis were obtained using loading projection of original variables. Next, we compared group performance after removing outliers at SD ≥ 3 for each group, aiming to improve the quality of the analysis. For a detailed explanation, refer to Online Resources 2.1.

**Figure 1.**
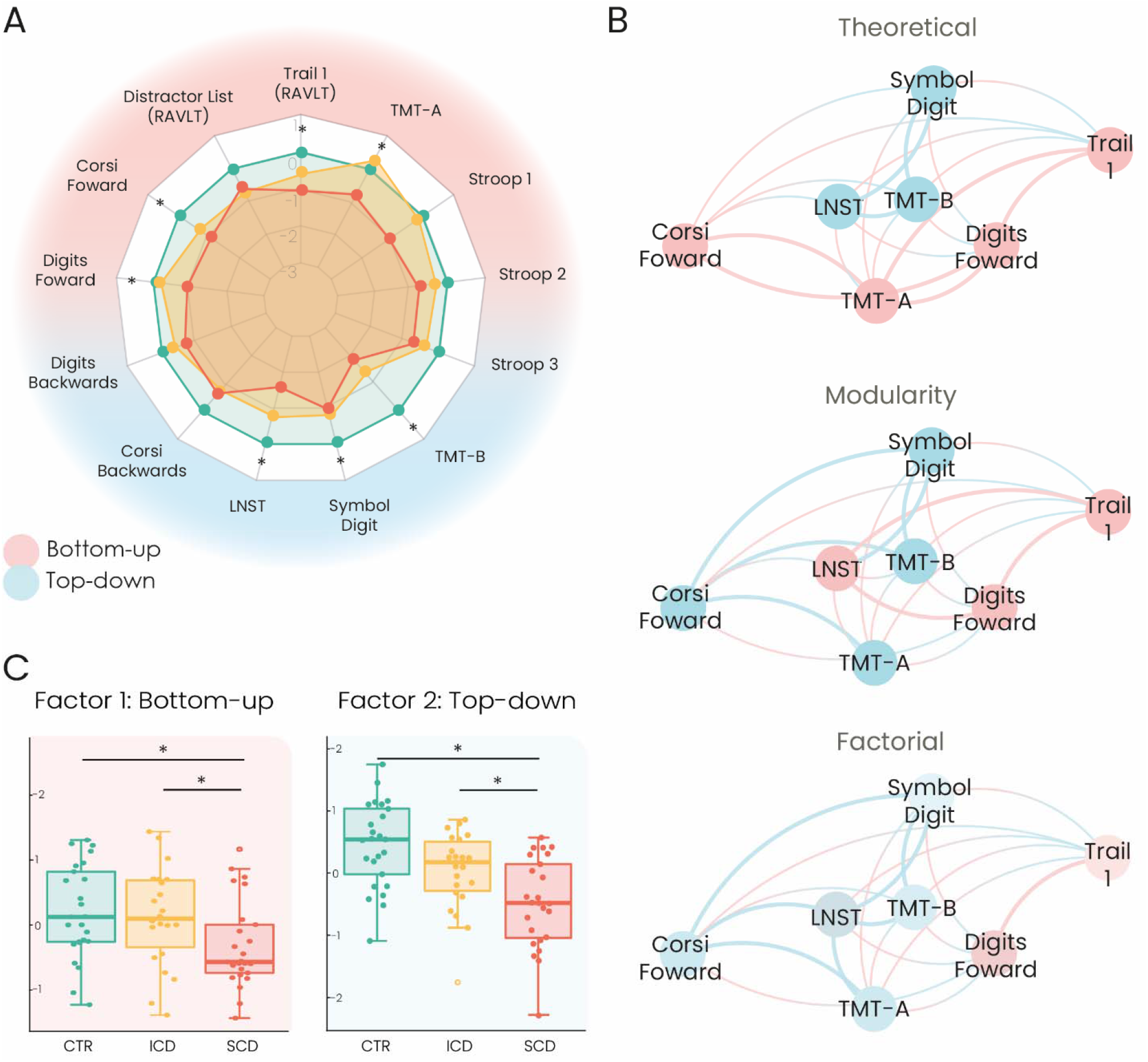
Behavioral performance: A. Comparison of the performance of each group across neuropsychological tasks. The performance of the SCD (red) and ICD (yellow) groups is compared to the reference values for the CTR group (green). Statistical significance was determined using Kruskal-Wallis tests between groups. B. Graphs of task performance similarity, colored by domain (top), community structure (mid) and factorial analysis (bottom). In each of these graphs, the nodes represent a single task and the edges connecting them represent the correlation between the associated performances (i.e. the closer the nodes, the higher the correlation). C. Factor analysis of neuropsychological measures. The 2 factors (Bottom-up, Top-down) are named based on their factor load in individual tasks within both domains. * indicate significant differences (*p*-FDR corrected <0.05). RAVLT: Rey Auditory Verbal Learning Test; TMT-A: Trail Making Test A; TMT-B: Trail Making Test B; LNST: Letter-Number Sequencing Test; CTR: Control; SCD: Smoked cocaine dependent; ICD: insufflated cocaine dependent.

#### 2.5.2 ERP analysis

ERP response between groups was compared using a point-by-point Monte Carlo permutation test with a significance level of *p* < .05 and a minimum cluster extension of five consecutive points, as in previous studies (de la Fuente et al., 2019; García-Cordero et al., 2016). As reported previously (Bekinschtein et al., 2009; Gonzalez-Gadea et al., 2015), MMN and P3a were analyzed from local deviant conditions from 100 to 200 ms and 200 to 400 ms, respectively; while the P3b was analyzed from global deviant conditions in two windows of analysis (whole ERP from 200 to 500 ms and maximum amplitude from 300 to 400 ms) on a frontal region of interest encompassing four electrodes (C25-C21-C12-C22 or Fz). The area over the curve (AOC) for P3a and P3b were used as representative measures of ERP modulation (for a detailed explanation, see Online Resources 4.1). Correlation analysis between ERP and cognitive data was performed by the Spearman correlation, considered significant at *p* < .05. Data analysis was conducted in R (version 4.1.2).

## 3. Results

### 3.1 Attentional cognitive tasks

SCD showed decreased performance with respect to CTR in Digits Forward, Corsi Forward, TMT-A, TMT-B, Symbol Digit, LSNT, and Trail 1 of RAVLT (*p* < .05, Fig. 1A). Also, SCD presented a reduced performance when compared to ICD in Digits Forwards and TMT-A (*p* < .05, Fig. 1A), while ICD did not differ significantly to CTR in any attentional test. See Online Resources (Table 5) and our previous article (de la Fuente et al., 2021) for further detail of the neuropsychological assessment and performance across multiple domains.

Following, we used the correlations of test performances to construct a graph representation of the tasks that yielded a significant difference between consumers and controls (see section 2.5.1 and Fig. 1B). This analysis allows for the exploration of communities formed by performance similarities which may match ad hoc grouping of the tests or bring up underlying structures based on performance. The graph, illustrated in Fig 1B (mid), presented an optimal modularity of 0.006, with 2 modules representing 31% and 18% of all tasks, respectively. According to the optimal modular structure and theoretical composition, an exploratory factor analysis with two factors was run (Table 2). Figure 1B shows graphs colored according to attentional domains (top), modularity (mid), and factorial analysis (bottom). Consistently with the theoretical and modularity structure, the factors reflected bottom-up and top-down processes similarly.

Finally, when comparing the performance of groups in the behavioral factors (Fig. 1C, further detail in Online Resources Table 6), we found that SCD performed poorer than both CTR and ICD in both factors (Bottom-up: H_(2, 72)_ = 7.97, *p* = .01, SCD vs. CTR *p* = .02, SCD vs. ICD *p* = .03; Top-down: H_(2, 72)_ = 17.96, *p* < .001, SCD vs. CTR *p* < .001, SCD vs. ICD *p* = .03). Although ICD did not differ from CTR, there is a tendency toward a decreased performance in the top-down factor than in the bottom-up (Bottom-up ICD vs. CTR *p* = .77; Top-down ICD vs. CTR *p* = .06).

### 3.2 Attentional ERP analysis

We performed this analysis on subjects performing the Local-Global Task, which is illustrated in Fig. 2A. All groups showed attentional ERP modulations (Online Resources, Fig. S1) with no measurable differences in data quality (Online Resources, Table 7) or baseline conditions between the groups (Online Resources, Fig. S2). Both groups of consumers exhibited a reduced modulation of the P3a (Fig. 2B, left) and P3b (Fig. 2B, right) when compared to CTR (5000 permutations, *p* < .05), while there were no significant differences in the MMN (*p* > .05). For the P3a response, the cluster of significant differences between SCD and CTR (t = 301.6 - 399.2 ms) was 2.78 times longer (62.5 ms) than that of ICD and CTR (t = 313.3 - 348.4 ms), with no differences in latency (H _(2, 72)_ = 1.07, *p* = .58). Similarly, for the P3b response, the cluster of significant differences between ICD and CTR (t = 317.2 – 340.6 & 356.3 – 399.2 ms) was 1.7 times longer (27.2 ms) than that of SCD and CTR (t = 352.3 – 391.4 ms), with no differences in latency (H _(72, 2)_ = 0.05, *p* = .97). These results indicate that the differences in the P3a were more prolonged in time for SCD, while in the P3b for ICD (Fig. 2B).

**Fig 2.**
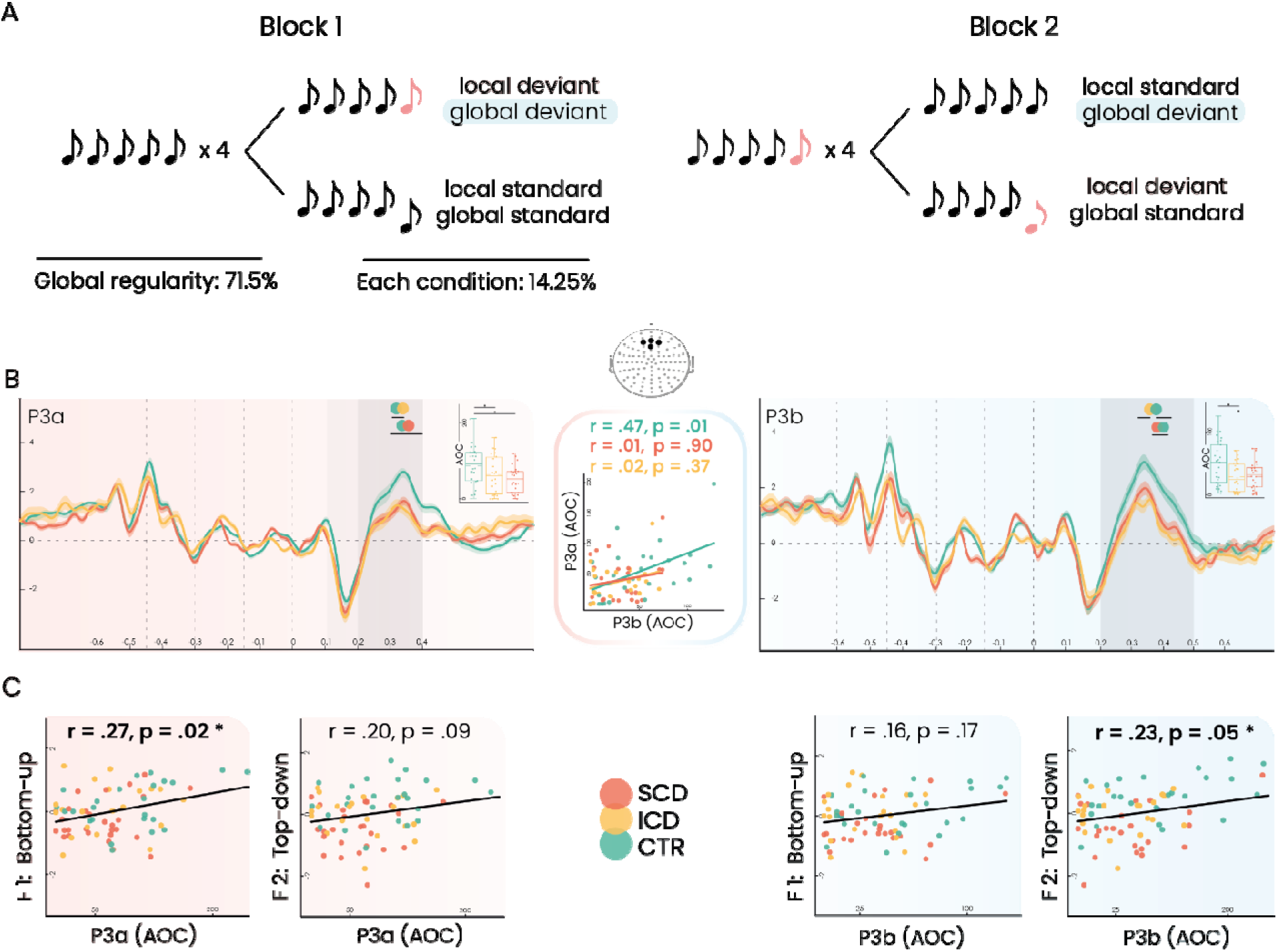
ERP analysis. A. Global-Local Task. Auditory stimuli consisted of 5 tones type A (black) or B (pink) presented in 2 blocks. B. ERPs response, P3a and MMN (left) and P3b (right) in the ROI. All differences reported were calculated via a point-by-point Monte Carlo permutation analysis (5000 permutations, *p* < .05). Shadowed lines indicate SEM. The gray box indicates the area of analysis. Black lines indicate the cluster of significant differences and the circles and asterisk indicate the groups differing. The scatterplot in the middle shows the association between P3a and P3b AOC by group. C. Scatterplots show the correlation between both factors (Bottom-up and Top-down) and ERP measures (AOC of P3a and AOC of P3b). ld: local deviant; ls: local standard; gd: global deviant; gs: global standard; Int: interaural; AOC: area over the curve; CTR: Control; SCD: smoked cocaine dependent; ICD: insufflated cocaine dependent.

We found a significant association between the AOC of P3a and P3b (r = .33, *p* = .005) across samples that remain significant and stronger per groups only for CTRs (r = .47, *p* = .01; SCD: r = .01, *p* = .9; ICD: r = .02, *p* = .37), as observed in Figure 2B (mid). Lastly, the ERPs were associated with the behavioral factors as expected (Fig. 2C): while the P3a was significantly associated with the bottom-up factor (r = .27, *p* = .02), the P3b was significantly associated with the top-down factor (r = .23, *p* = .05). There were no significant associations in the remaining two combinations (P3a with top-down factor, and P3b with bottom-up factor). These associations became stronger only for CTR, although they did not reach the significant threshold (P3a and Bottom-up: CTR r = .35, *p* = .08, ICD r = .22, *p* = .30, SCD r = .06, *p* = .7; P3b and Top-down: CTR r = .31, *p* = .12, ICD r = .12, *p* = .5, SCD r = .07, *p* = .7).

## 4. Discussion

Although previous studies have identified deficits in bottom-up/P3a and top-down/P3b among cocaine consumers, no study has explored the impact of the route of administration (Anderson et al., 2011; Bauer, 1997; Biggins et al., 1997; Gooding et al., 2008; Habelt et al., 2020; Houston and Schlienz, 2018; Wakim et al., 2021). Our study presents unprecedented evidence revealing distinct modulations of attentional ERPs based on the routes of administration. Our findings align with prior research indicating larger attentional deficits in SCD compared to CTR and ICD (de la Fuente et al., 2021; Oliveira et al., 2018), both at the neurophysiological and neuropsychological levels. Specifically, we observed pronounced impairments in the P3a component for SCD, along with greater attentional deficits in cognitive tasks. Moreover, we found more extensive differences in the P3b component for ICD. These results may contribute to the development of interventions targeted at specific attentional processes as well as a better understanding of the underlying mechanism of these deficits.

Regarding attentional neurocognitive performance, SCD displayed reduced values compared to CTR and ICD (see Table S5). The performance correlations graph modularity aligned with the theoretically expected, although Corsi Forward, TMT-A exhibited higher load on the top-down factor. This outcome is reasonable considering that cognitive tasks inherently involve varying degrees of executive functions, memory, and other cognitive domains (Goldstein et al., 2004), making it challenging to separate them into distinct factors. Particularly, Corsi Forward and TMT-A involve various complex cognitive functions, including visuospatial skills (Corsi, 1972; Reitan and Wolfson, 1993). SCD exhibited reduced performance in both bottom-up and top-down processes compared to CTR and ICD. This finding aligns with previous studies that reported more pronounced attentional impairments for faster routes of administration (de la Fuente et al., 2021; Oliveira et al., 2018). While ICD did not significantly differ from CTR, there was a noticeable trend toward top-down impairment (*p* = .06), in agreement with research indicating impaired memory and executive function but not attentional deficits in ICD (de la Fuente et al., 2021; Oliveira et al., 2018).

### 4.1 Electrophysiological markers of attention

At a pre-attentional level, we found no significant differences in MMN modulation between the groups, indicating that early stages of auditory processing are preserved in cocaine consumers regardless of the administration route. In line with previous studies (Anderson et al., 2011; Bauer, 1997; Habelt et al., 2020; Houston and Schlienz, 2018; Wakim et al., 2021), we found lower P3a and P3b responses in both groups of consumers. Our findings suggest that these alterations are common across different routes of administration, and they could either be associated with drug use (Hirsiger et al., 2019; Vonmoos et al., 2014) or show a vulnerability towards consumption (Harper et al., 2021; Tervo-Clemmens et al., 2018).

Regarding bottom-up attentional processing, SCD exhibited larger differences in the P3a, consistent with research at neurochemical, connectivity and cognitive levels. First, P3a relies on tonic dopamine levels (Heitland et al., 2013; Kähkönen et al., 2002; Rangel-Gomez et al., 2013), critical for attentional capture (Corbetta et al., 2008). SC has a strong impact on the reward system, leading to an increase in phasic dopamine in the short term and a decrease in tonic dopamine in the long term (Nieoullon, 2002; Samaha et al., 2004; Samaha and Robinson, 2005). Second, the frontal regions involved in the P3a/bottom-up (Fjell et al., 2005; Friedman et al., 2001; Polich, 2007) exhibit disrupted connectivity in SCD (de la Fuente et al., 2021; Oliveira et al., 2018; Rosário et al., 2019). Altogether, these findings suggest that local processes related to attentional capture are more compromised in faster routes of administration. Even though ICD also presented lower P3a modulation compared to CTR, these differences were less extensive than those for SCD, and were not present at bottom-up cognitive level. Attentional deficits are a distinct signature of SCD (de la Fuente et al., 2021), and the lack of cognitive differences accompanying lower P3a modulation in ICD may be due to the partial reversibility of drug-related cognitive deficits after abstinence (Hirsiger et al., 2019; Vonmoos et al., 2014) and the plasticity of adolescent brain (Grant and Dawson, 1998).

Regarding top-down processing, ICD exhibited more prolonged differences in the P3b, consistent with research at anatomical and cognitive levels. P3b is linked to memory processing in parietal regions (Polich, 2007) and ICD exhibit lower gray matter density over parietal areas compared to SCD as well as executive and memory impairments (de la Fuente et al., 2021). These findings suggest slower routes of administration primarily affect sustaining attention, which in turn may explain previous contradictory findings (Campanella et al., 2019; Houston and Schlienz, 2018). Along with the non-significant differences in top-down performance with CTR, these results point toward a reduced affectation for slower routes compared to faster routes, consistent with previous research (Allain et al., 2015; Castaño, 2000; de la Fuente et al., 2021; Meyer and Quenzer, 2019; Minogianis et al., 2013; Oliveira et al., 2018; Samaha and Robinson, 2005). These distinct results for SC and IC highlight the significance of exploring the administration route.

Finally, the association between P3a and P3b modulation was observed in CTR but not in consumers, regardless of the route of administration. This has two main theoretical and clinical implications. Theoretically, our electrophysiological findings support the relationship between top down and bottom-up processes. Conflicting findings exist regarding the association or independence of these processes (Buschman and Miller, 2007; Karakaş et al., 2003; Leij et al., 2013; Schneider et al., 2014). On one hand, at the genetic and behavioral level, these processes seem independent, as different genetic alleles relate exclusively to each process and there are noticeable differences in cognitive performance (Leij et al., 2013; Schneider et al., 2014). On the other hand, anatomical studies suggest a relationship (Buschman and Miller, 2007; Katsuki and Constantinidis, 2014; Li et al., 2013; Posner and Petersen, 1990; Yamaguchi et al., 2000), particularly in the auditory domain (Alho et al., 2015). Furthermore, each factor correlated significantly with its corresponding ERP measure (P3a for bottom-up and P3b for top-down), supporting their reliability as attentional biomarkers. Clinically, the absence of this association in consumers is consistent with previous literature (Ma et al., 2014; Moeller et al., 2010; Orsini et al., 2018; Tomasi et al., 2007; Zhai et al., 2022). Although our cross-sectional study cannot identify vulnerability or drug-derived impairment, these results are clinically relevant for the cognitive treatment of patients with cocaine addiction. This lack of association is consistent with their lower modulation of ERPs and cognitive performance. This may be attributed to alterations in the prefrontal and parietal cortex among cocaine consumers (Ma et al., 2014; Moeller et al., 2010; Orsini et al., 2018; Tomasi et al., 2007; Zhai et al., 2022), which play a crucial role in bottom-up and top-down processes (Buschman and Miller, 2007; Katsuki and Constantinidis, 2014; Li et al., 2013; Posner and Petersen, 1990; Yamaguchi et al., 2000).

### 4.2 Limitations and further directions

The first limitation to this study is that the sample size was rather small, but we addressed this by controlling for demographic, clinical, and poly-consumption variables (Online Resources 1), which were not adequately controlled for in previous research. Consequently, the consumers differed only in the route of administration (SC or IC), a relevant factor that is frequently overlooked in the literature (Frazer et al., 2018; Goldstein et al., 2004; Gooding et al., 2008; Lopes et al., 2017; Potvin et al., 2014; Vicario et al., 2020; Vonmoos et al., 2013; Wakim et al., 2021). Second, we had a larger proportion of male participants, although this reflects that the sample was drawn from a real-world clinical population as drug abuse incidence is higher among males (Ángeles, 2013). Third, using ERPs has limitations due to their high variability, however, our findings are supported by converging behavioral and neuropsychological data. Fourth, the cognitive tasks used were not specifically designed to measure bottom-up and top-down processes, although some have been previously used for this purpose (Araneda et al., 2015; Gevers et al., 2015; Schneider et al., 2014). Future studies could employ tasks designed for these processes (e.g., Posner, 1980). Lastly, our study is cross-sectional, highlighting the need for future longitudinal studies to explore the relationship between neurocognitive deficits and cocaine intake.

## 5. Conclusions

Our results cast a new light on decreased attentional ERP modulation based on the route of cocaine administration, finding subtle yet significant differences. SCD showed greater attentional impairment, mainly at bottom-up/P3a processes, while ICD showed a trend towards top-down/P3b deficits. This indicates that faster routes of administration are associated with larger attentional capture deficits, while slower routes are linked to difficulties in sustaining attention. Longitudinal research studies suggest a bidirectional relationship between attention deficits and substance use (Harper et al., 2021; Hirsiger et al., 2019; Tervo-Clemmens et al., 2018; Vonmoos et al., 2014) and our findings suggest that the route of administration may play a key role in determining the severity of these deficits. These findings highlight the importance of considering the route of administration of stimulant drugs in both clinical and research settings. The route of cocaine administration is linked to distinct neurocognitive profiles, and our study yields translational results that align neurophysiological and neuropsychological levels. Furthermore, our findings strengthen the validity of utilizing attentional ERPs (P3a and P3b) as reliable measures for assessing attentional deficits in the context of substance abuse.

## Data Availability

All data produced in the present work are contained in the manuscript

## Founding

This work was supported by the Florencio Perez Foundation and the INECO Foundation.

## Author contributions

Agustina Aragón-Daud: Conceptualization, Data curation, Formal analysis, Investigation, Methodology, Project administration, Visualization, Writing -Original Draft and review & editing. Alethia de la Fuente: Conceptualization, Data curation, Formal analysis, Investigation, Methodology, Project administration, Visualization, Writing -Original Draft and review & editing, Supervision. Sofía Schurmann Vignaga, Sofía Milagros Oberti de Luca, Pilar Prado, Rosario Figueras and Lucia Lizaso: Data curation. Facundo Manes: Conceptualization, Investigation, Funding acquisition, Writing - review & editing. Marcelo Cetkovich: Conceptualization, Investigation, Writing - Original Draft and review & editing. Teresa Torralva: Conceptualization, Project administration, Methodology, Supervision, Visualization, Writing - review & editing.

All authors have approved the final article.

## Acknowledgments

The authors acknowledge the Federación de Organizaciones no Gubernamentales de la Argentina para la Prevención y el Tratamiento de Abuso de Drogas (FONGA) as well as the patients, clinicians, and operators of the therapeutic communities (Buen Sanmaritano, El Reparo, Foundation Creer es crear, Creando la libertad, El Palomar, Modelo Minnesota). In addition, we acknowledge the generosity of the non-profit organizations that contributed to the recruitment of controls (Center Juan Pablo II, Foundation Temas, Matanza secretary, Agustin at Barrio Mitre).

